# Double-zero-event studies matter: a re-evaluation of physical distancing, face masks, and eye protection for preventing person-to-person transmission of COVID-19 and its policy impact

**DOI:** 10.1101/2020.08.12.20173674

**Authors:** Mengli Xiao, Lifeng Lin, James S. Hodges, Chang Xu, Haitao Chu

**Affiliations:** Division of Biostatistics, School of Public Health, University of Minnesota, Minneapolis, MN, USA; Department of Statistics, Florida State University, Tallahassee, FL, USA; Department of Population Medicine, College of Medicine, Qatar University, Doha, Qatar

**Keywords:** COVID-19, double-zero-event study, meta-analysis, policy making, sensitivity analysis

## Abstract

**Objectives:** High-quality meta-analyses on COVID-19 are in urgent demand for evidence-based decision making. However, conventional approaches exclude double-zero-event studies (DZS) from meta-analyses. We assessed whether including such studies impacts the conclusions in a recent systematic urgent review on prevention measures for preventing person-to-person transmission of COVID-19.

**Study designs and settings:** We extracted data for meta-analyses containing DZS from a recent review that assessed the effects of physical distancing, face masks, and eye protection for preventing person-to-person transmission. A bivariate generalized linear mixed model was used to re-do the meta-analyses with DZS included. We compared the synthesized relative risks (RRs) of the three prevention measures, their 95% confidence intervals (CI), and significance tests (at the level of 0.05) including and excluding DZS.

**Results:** The re-analyzed COVID-19 data containing DZS involved a total of 1,784 participants who were not considered in the original review. Including DZS noticeably changed the synthesized RRs and 95% CIs of several interventions. For the meta-analysis of the effect of physical distancing, the RR of COVID-19 decreased from 0.15 (95% CI, 0.03 to 0.73) to 0.07 (95% CI, 0.01 to 0.98). For several meta-analyses, the statistical significance of the synthesized RR was changed. The RR of eye protection with a physical distance of 2 m and the RR of physical distancing when using N95 respirators were no longer statistically significant after including DZS.

**Conclusions:** DZS may contain useful information. Sensitivity analyses that include DZS in meta-analysis are recommended.

## 1. Introduction

As of August 3, 2020, the worldwide daily number of new cases of coronavirus disease 2019 (COVID-19) is more than 2 million, with over 6.9 million deaths documented so far.^1^ Concerns about reopening schools are front-page news as fall semester approaches.^2-5^ High-quality evidence is in urgent demand for guiding educational decision makers regarding reopening schools.^2,4^ Without proven vaccines or therapies, decisions on school reopening can rely only on behavior-based preventions to contain the spread of COVID-19.^6^ However, evidence about prevention measures appears heterogeneous and is inconsistently interpreted by policy makers.^7-9^ The US Centers for Disease Control and Prevention, for example, shifted to advocate the use of face masks from previous advice against wearing face masks in public.^10^ Questions still remain about what levels of prevention measures and their interactions can best protect people from COVID-19 infections.

Systematic reviews and meta-analyses can offer high-quality evidence by synthesizing all available, though possibly inconsistent, studies about the effects of prevention measures. Recently, Chu et al.^9^ performed a systematic review on the effectiveness of face masks, eye protection, and physical distancing for preventing COVID-19, providing important evidence to policymakers in this fast-evolving situation. The reviewers identified 44 observational studies of severe acute respiratory syndrome (SARS), Middle East respiratory syndrome (MERS), and COVID-19. These studies’ results were synthesized in multiple meta-analyses based on virus types, prevention measures, and healthcare settings. These meta-analyses found reduced risks of infection with face masks, eye protection, and physical distancing.

Because the sample sizes in many studies, especially those on COVID-19, are not large, the systematic review contains a considerable number of studies with zero counts of infection events, creating challenges in estimating effect sizes. If a zero count appears only in a single intervention arm, a fixed value of 0.5 is conventionally added to all four data cells in the 2×2 table of the associated study.^11^ If zero counts appear in both arms, this double-zero-event study (DZS) is conventionally omitted from the analyses. Among the 44 studies in this review, at least 9 contain DZS. The meta-analyses in Chu et al.^9^ excluded these DZS, as implied in the associated forest plots, and thus lost information about a total of 1,784 subjects.

Although artificial correction and omission of zero event counts are default procedures for DZS in most meta-analysis software, these methods can impact the meta-analytic conclusions. The statistical significance, the direction of intervention effect, or both, can change after excluding DZS based on both simulation studies and empirical data analyses.^12–15^ Multiple simulation studies have demonstrated that effect estimates could be biased with poor coverage probability when artificially correcting or omitting zero event counts.^12,15–17^ For example, the clinically relevant advantage of off-pump surgery for coronary artery bypass grafting became statistically significant after including DZS.^17^ In another study re-analyzing 442 meta-analyses from the *Cochrane Database of Systematic Reviews*, 41 meta-analyses changed statistical significance and the effect direction was reversed in 8 meta-analyses after including DZS.^15^

Several statistical approaches can be used to account for DZS in meta-analyses without artificial correction or omission of zero event counts. For example, one may use the so-called confidence distribution for hypothesis testing to obtain point estimates for DZS.^18^ Using this confidence distribution approach to combine all studies, including DZS, Liu et al.^18^ found that the effect of rosiglitazone on myocardial infarction had a narrower confidence interval (CI) than that produced by the conventional method excluding DZS. However, this approach needs complicated procedures for implementation and may not be intuitive for clinical applications. Alternatively, generalized linear mixed models can be also used to include zero event counts in meta-analysis without laborious computational procedures.^15,19,20^ They directly model event counts within studies and account for the correlation of effect sizes between the intervention and control groups.^20,21^

This article aims at assessing the impact of DZS on meta-analysis results about prevention measures for COVID-19 and their reliability for policy-making. We use a bivariate generalized linear mixed model (BGLMM) to reanalyze the meta-analyses presented in Chu et al.,^9^ accounting for all DZS, and we re-evaluate the conclusions from that systematic review.

## 2. Methods

The BGLMM for a meta-analysis of prevention measures for COVID-19 can be specified as follows. Let *n_i_*_0_ be the number of participants in the control (i.e., no prevention) group, and *n_i_*_1_ be the number of participants in the intervention (i.e., prevention) group in the ith study, for *i =* 1,…, *N*, where *N* is the number of studies in the meta-analysis. Moreover, in the ith study, denote the underlying true probabilities of COVID-19 infection events in the control and prevention groups as *p_i_*_0_ and *p_i_*_1_, respectively. Let the event counts be *y_i_*_0_ for the control group and *y_i_*_1_ for the prevention group. The event counts *y_i_*_0_ and *y_i_*_1_ follow a binomial distribution, and they are independent within and across studies:

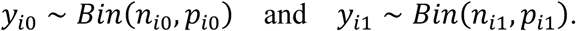

In conventional meta-analyses, the event probabilities are estimated as 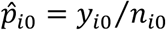 and 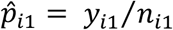; both 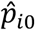 and 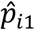 are zero for DZS studies (i.e., *y_i_*_0_ *= y_i_*_1_ *=* 0), which create difficulty in estimating the effect sizes in such studies.

Instead of estimating the effect sizes of individual studies, the BGLMM directly models the event counts *y_i_*_0_ and *y_i_*_1_ with binomial likelihoods, so that zero event counts are permitted. Although the event counts are independent within studies because they are based on different groups of participants, the underlying true event probabilities *p_i_*_0_ and *p_i_*_1_ are potentially correlated. Specifically, they can be modeled jointly as:

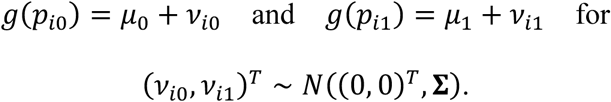

Here, *g(·)* denotes the link function that transforms the event probabilities to linear forms. The parameters *μ*_0_ and *μ*_1_ are fixed effects, representing the overall event probabilities on the transformed scale. The study-specific parameters *ν_i_*_0_ and *ν_i_*_1_ are random effects and assumed to follow a bivariate normal distribution with zero means and variance-covariance matrix

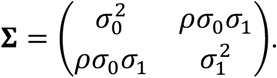

The parameters 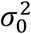 and 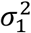 are between-study variances for the two groups due to heterogeneity, and *ρ* is the correlation between the two groups.

This article uses the probit link; it leads to closed forms for the marginal event probabilities 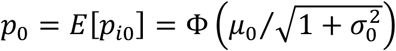 and 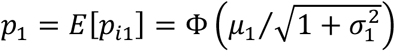. We focus on estimating the overall relative risk (RR), which is the effect measure in the original meta-analyses on COVID-19. The overall RR is *p*_1_*/p*_0_. We obtain the standard error of the estimated overall RR, and thus the Wald-type CI, on a logarithmic scale, and then back-transform the CI to the scale of RR.

Of note, other link functions (e.g., the logit link) may be also used in the BGLMM above, although generally they do not lead to closed-forms for the marginal event probabilities, which need to be obtained via approximations. Nevertheless, because the probit and logit functions are fairly close, they are expected to produce similar results. Also, once we estimate the marginal event probabilities *p*_0_ and *p*_1_, it is also easy to obtain other summary effect measures (e.g., odds ratio and risk difference).

We applied BGLMMs to re-analyze the meta-analyses containing zero counts that were presented in Chu et al.^9^ Specifically, these meta-analyses correspond to their Figures 2, 4, 6, and the third and fourth subfigures in Appendix 6, which respectively investigated the risk with physical distancing (categorized by virus types), face masks (categorized by healthcare settings), eye protection (categorized by virus types), eye protection comparing different physical distances, and physical distancing for different face masks. The BGLMMs were implemented using PROC NLMIXED in SAS (version 9.4). The random effects were approximately integrated by the default adaptive Gaussian quadrature, and likelihood maximization used the default dual quasi-Newton optimization algorithm.

## 3. Results

Table 1 compares the results of our re-analyses including DZS with those excluding DZS; Figure 1 visualizes these results. Including DZS noticeably changed the synthesized RRs and their 95% CIs of several interventions. Some RRs shifted towards the null value 1, while others became smaller or more evident. For example, among the meta-analyses of physical distancing, the RR in the SARS subgroup increased from 0.35 (95% CI, 0.23 to 0.52) to 0.40 (95% CI, 0.27 to 0.58) after including DZS. The RR in the MERS subgroup decreased from 0.23 (95% CI, 0.04 to 1.20) to 0.18 (95% CI, 0.03 to 0.99), and in the COVID-19 subgroup decreased from 0.15 (95% CI, 0.03 to 0.73) to 0.07 (95% CI, 0.01 to 0.98). Including DZS also changed the statistical significance of several RRs. For MERS, the RR of physical distancing was non-significant when excluding DZS but became significant after including DZS; in contrast, the RR of eye protection changed from significant to non-significant.

**Table 1.** Comparison between the synthesized intervention effects with and without double-zero-event studies in meta-analyses on physical distancing, face masks, and eye protection

| Meta-analysis | No. of subjects in DZS (percentage of total sample size) | No. of DZS (percentage of all studies) | RR (95% CI) with DZS excluded | RR (95% CI) with DZS included |
| --- | --- | --- | --- | --- |
| **Risk with physical distancing** |  |  |  |  |
| MERS (n=1158, N=8) | 201 (17.4%) | 4 (50.0%) | 0.23 (0.04, 1.20) | 0.18 (0.03, 0.99) |
| SARS (n=9101, N=17) | 41 (0.5%) | 1 (5.9%) | 0.35 (0.23, 0.52) | 0.40 (0.27, 0.58) |
| COVID-19 (n=477, N=7) | 204 (42.8%) | 2 (28.6%) | 0.15 (0.03, 0.73) | 0.07 (0.01, 0.98) |
| Overall (n=10736, N=32) | 446 (4.2%) | 7 (21.9%) | 0.30 (0.20, 0.44) | 0.32 (0.21, 0.49) |
| **Risk with face masks** |  |  |  |  |
| Healthcare setting (n=9445, N=26) | 358 (3.8%) | 6 (23.1%) | 0.30 (0.22, 0.41) | 0.34 (0.23, 0.51) |
| Overall (n=10170, N=29) | 358 (3.5%) | 6 (20.7%) | 0.34 (0.26, 0.45) | 0.37 (0.25, 0.54) |
| **Risk with eye protection** |  |  |  |  |
| MERS (n=1056, N=4) | 34 (3.2%) | 1 (25.0%) | 0.24 (0.06, 0.99) | 0.22 (0.01, 4.41) |
| SARS (n=2581, N=8) | 134 (5.2%) | 2 (25.0%) | 0.34 (0.21, 0.56) | 0.36 (0.24, 0.55) |
| Overall (n=3713, N=13) | 244 (6.6%) | 4 (30.8%) | 0.34 (0.22, 0.52) | 0.35 (0.21, 0.59) |
| **Risk with eye protection comparing different physical distances** |  |  |  |  |
| 0 m (n=881, N=4) | 102 (11.6%) | 1 (25.0%) | 0.21 (0.13, 0.34) | 0.23 (0.15, 0.32) |
| 1 m (n=1786, N=6) | 118 (6.6%) | 3 (50.0%) | 0.53 (0.39, 0.72) | 0.55 (0.35, 0.85) |
| 2 m (n=1084, N=5) | 62 (5.7%) | 2 (40.0%) | 0.24 (0.06, 0.99) | 0.21 (0.01, 7.09) |
| Overall (n=3756, N=15) | 288 (7.7%) | 6 (40.0%) | 0.34 (0.22, 0.52) | 0.35 (0.21, 0.50) |
| **Risk with physical distancing for different face masks** |  |  |  |  |
| N95 respirators (n=1213, N=12) | 370 (30.5%) | 6 (50.0%) | 0.38 (0.16, 0.91) | 0.51 (0.22, 1.15) |
| All non-N95 masks (n=9531, N=20) | 78 (0.8%) | 1 (5.0%) | 0.27 (0.17, 0.43) | 0.27 (0.16, 0.46) |
| Overall (n=10744, N=32) | 448 (4.2%) | 7 (21.9%) | 0.30 (0.20, 0.44) | 0.33 (0.22, 0.50) |

n=number of subjects. N=number of studies. SARS=severe acute respiratory syndrome. MERS=Middle East respiratory syndrome. RR=relative risk. DZS=double-zero-event studies. CI=confidence interval.

**Figure 1.**
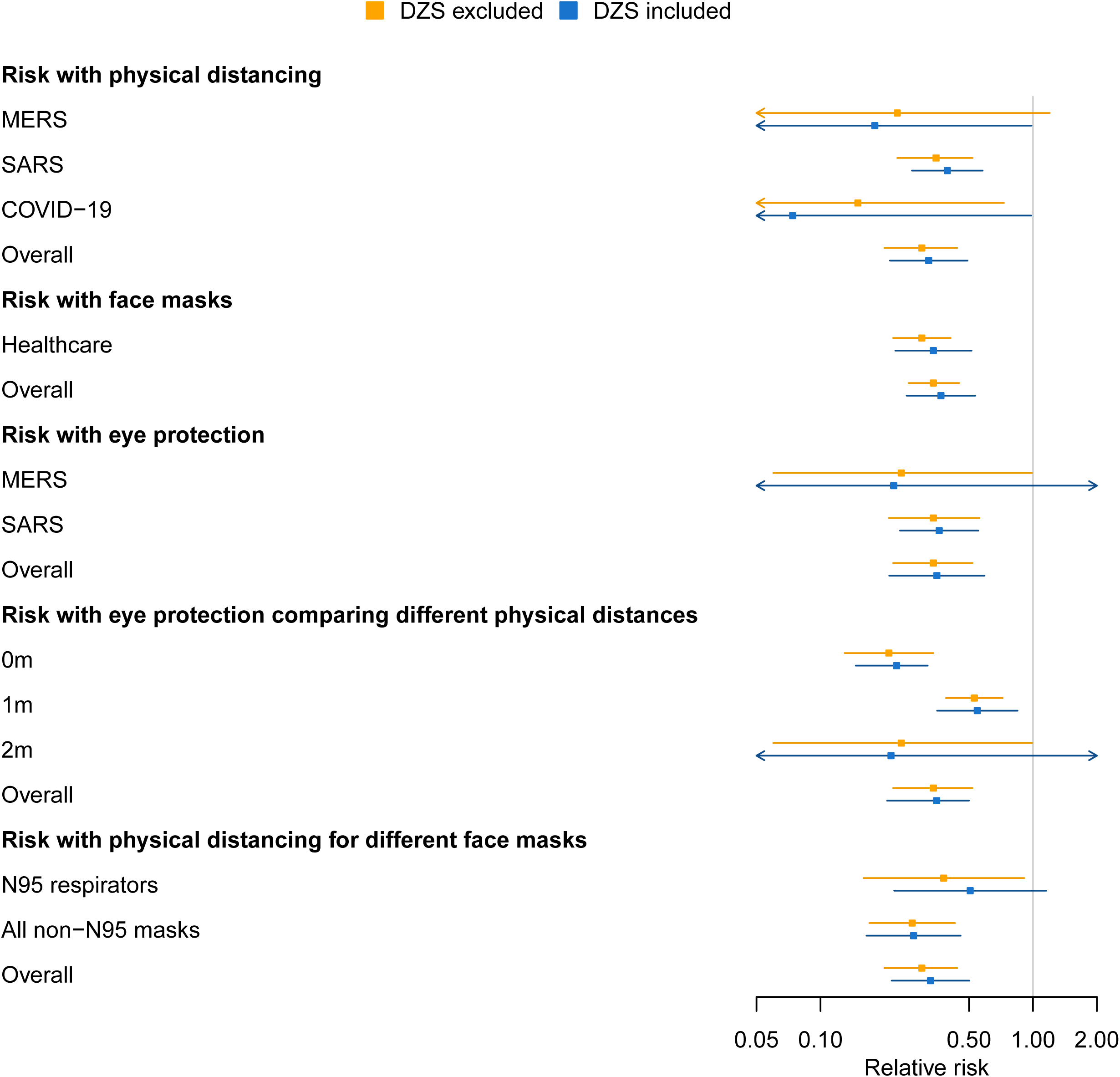
Synthesized relative risks and 95% confidence intervals with double-zero-event studies excluded and included in meta-analyses on prevention measures for COVID-19

These changes were related to the proportions and sample sizes of DZS in the corresponding meta-analyses (Table 1). In the MERS subgroup, 4 out of 8 (50%) studies were DZS, containing 201 of 1158 subjects. In the COVID-19 subgroup, 2 out of 7 (29%) studies were DZS, containing 204 of 477 subjects. The DZS contained considerable information in these subgroups; thus, the RRs produced by excluding and including DZS had noticeable differences.

Such a trend was also observed when including DZS in the two meta-analyses that focused on combinations of different prevention approaches. By including DZS, our re-analyses show that with physical distance of 2 m, the effect of eye protection on reducing infection risk was no longer statistically significant, and the reduced risk of physical distancing might not be statistically significant when using N95 respirators.

## 4. Discussion

This article re-evaluated the evidence about measures for preventing person-to-person transmission of COVID-19, including physical distancing, face masks, and eye protection, in a recent timely systemic review by accounting for DZS in meta-analyses.^9^ We found that including DZS provided useful information about prevention of COVID-19. Although most results were consistent with those in the original review that omitted DZS from meta-analyses, including DZS turned the significant effect of eye protection to non-significant when physical distancing is 2 m. Including DZS also implied that the effect of physical distancing was no longer statistically significant when using N95 respirators, while it did not change the conclusion for non-N95 face masks.

Given that high-quality evidence about COVID-19 is in great demand and some intervention effects can be changed by including DZS, we suggest future systematic reviews and meta-analyses on COVID-19 properly consider the impact of including DZS. When policymakers are weighing the public health risks of COVID-19 about reopening schools against economic and educational loss, reliable evidence is urgently needed, while early-stage studies on COVID-19 may have small sample sizes and thus zero event counts. Sensitivity analyses that include important information from DZS are recommended to validate evidence from meta-analyses. If including DZS does not substantially change the conclusions, the meta-analyses can be used as reliable and robust evidence by policymakers.

The minimal physical distancing requirement varies in different countries from 1 m to 2 m, while the shortage of protective equipment (e.g., goggles and face shields) makes it impractical to recommend eye protection in all reopening schools. Our results show that 2 m physical distancing is noticeably more effective than 1 m and can potentially remedy the lack of eye protection. These findings suggest that education decision makers consider 2 m as a guideline for physical distancing when planning classroom capacity and seat adjustment, especially when eye protection is not widely available for students.

We also found no significantly reduced risk of physical distancing when using N95 respirators, while physical distancing still played an important role when using other types of face masks. This does not nullify the effect of physical distancing; rather, it implies that reopening schools may adapt their plans to different scenarios to effectively protect school personnel and students. For example, N95 respirators should be offered or prioritized when physical distancing is not possible; otherwise, use of normal face masks with strict physical distancing may be sufficient.

This article used the BGLMM to account for DZS in the meta-analyses of RRs; other approaches are also available to incorporate DZS, but some are only applicable to a specific effect measure, such as odds ratio or risk difference.^14,15,17,18,20^ Further work may evaluate whether other choices of methods or effect measures still support the existing evidence. Researchers who wish to include DZS in future meta-analyses can find our computer code in the Supplemental File.

## Data Availability

The data is published and available from the Lancet.

https://www.thelancet.com/journals/lancet/article/PIIS0140-6736(20)31142-9/fulltext

